# Impact of Polypharmacy on Medication Adherence in Patients with Comorbid Type 2 Diabetes and Hypertension in Tanzania: A Cross-Sectional Study

**DOI:** 10.64898/2026.08.14.26360430

**Authors:** Faidon B. Kihombo, Helfrid Ilomo, Martine A. Manguzu, Alphonce I. Marealle, Ritah F. Mutagonda

**Affiliations:** Department of Clinical Pharmacy and Pharmacology, School of Pharmacy, Muhimbili University of Health and Allied Sciences, P.O Box 65013, Dar es Salaam, Tanzania; Pharmacy Department, Muhimbili National Hospital, P.O Box 65000, Dar es Salaam. Tanzania

## Abstract

**Background:** Diabetes mellitus and hypertension are increasingly prevalent non-communicable diseases that often coexist due to their interrelated pathophysiology and commonalities of risk factors. Effective management of these two comorbid conditions often involves polypharmacy, defined as the concurrent use of five or more medications, which places a substantial pill burden on patients and demands consistent medication adherence for effective disease control. In Tanzania, where access to chronic care services remains constrained and out-of-pocket medication costs can be substantial, understanding the drivers of non-adherence is considered imperative.. Limited data exist on the extent of polypharmacy and its impact on adherence among Tanzanian patients with these comorbidities. This study therefore aimed at evaluating the prevalence of polypharmacy and its impact on medication medication adherence among this population.

**Methodology:** A cross-sectional study involving 396 outpatients was conducted at Muhimbili National Hospital. Consecutive sampling was used to recruit eligible participants. Data were collected using a structured questionnaire that captured information on socio-demographics, clinical characteristics, and adherence behaviors. Polypharmacy was defined as the concurrent use of five or more medications. Medication adherence was assessed using the Medication Adherence Report Scale (MARS-5). Modified Poisson regression with robust standard errors was performed to identify factors associated with medication adherence and to estimate crude and adjusted prevalence ratios (cPR and aPR) with 95% confidence intervals.

**Results:** 71% of the study participants were on five or more medications, indicating high polypharmacy prevalence, with a median of six medications. Medication adherence was reported at 55.1%. Among participants with lower adherence, a higher proportion had elevated fasting glucose for moderate (APR: 0.83, P = 0.001) and high fasting glucose (APR: 0.66, P < 0.001), and uncontrolled blood pressure while herbal medicine use (APR: 0.72, P < 0.001) was identified as contributing factor to lower adherence..

**Conclusion:** This study reveals a high prevalence of polypharmacy with moderate medication adherence among patients with comorbid T2DM and hypertension. These findings underscore the need for targeted interventions such as structured patient education, pharmacist-led medication reviews, and polypharmacy rationalization to improve medication adherence and clinical outcomes in this population.

## Introduction

Hypertension can be defined as having a systolic blood pressure(SBP) ≥140 mmHg or diastolic blood pressure(DBP) ≥90 mmHg, confirmed by at least two separate readings(1) whereas type 2 diabetes mellitus (T2DM) is a chronic metabolic disorder characterized by higher blood glucose levels that occurs when the body becomes resistant to insulin or does not make enough insulin. Over time, this increased glucose level leads to severe damage to many of the body’s systems particularly nerves and blood vessels(2).

Both hypertension and diabetes have a higher likelihood of co-existence due to the overlapping nature of their risk factors (i.e. physical inactivity, obesity and poor dietary practices). Additionally, these two conditions have a pathophysiological linkage in a way that diabetes results in microvascular complications that increase peripheral vascular resistance due to increase in vascular stiffness therefore leading into hypertension(3)

The global burden of hypertension and diabetes has risen substantially over the years, presenting a significant public health challenge. According to the International Diabetes Federation (IDF) atlas of 2025, an estimated 589 million people live with diabetes worldwide. This number is projected to rise to 643 million by 2030 and 783 million by 2045(2). In Africa, approximately 25 million adults have diabetes, this number is predicted to increase to 60 million by 2050 (3). On the other hand, WHO estimates that 1.28 billion adults aged 30 to 79 years have hypertension (WHO 2021).

Nearly two-thirds of hypertension and diabetes cases occur in low- and middle-income countries (LMICs), where health systems are already burdened with infectious diseases(3).East Africa alone has an estimated prevalence of 53% [95% CI: 45.8%–59.1%] of hypertension among people diagnosed with diabetes(3).Importantly, the co-occurrence of hypertension and diabetes is associated with a 44% increased risk of mortality and a 41% increased risk of cardiovascular events, compared to only 7% and 9% for diabetes alone, respectively (4).

Clinical management usually employs a multifactorial approach involving both non-pharmacological and pharmacological interventions. Non-pharmacological strategies focus on lifestyle modifications, including weight control, exercise, and dietary changes. However, pharmacological interventions are often necessary to meet treatment targets (5)and usually involve polypharmacy, defined as the concurrent use of multiple medications to control blood pressure, glucose levels, dyslipidemia, and associated pro-inflammatory and hypercoagulable states(6).

While polypharmacy can be beneficial, inappropriate medication use, drug interactions, adverse effects, and increased medication costs may negatively impact patient adherence to treatment regimens(7). Poor adherence increases the risk of recurrence and complications, adversely affecting quality of life, increasing healthcare costs, and elevating mortality rates(8). Therefore, maintaining adherence to medications is crucial for achieving treatment goals and improving outcomes in patients with comorbid hypertension and diabetes. Despite the rising burden of comorbid T2DM and hypertension in Tanzania, published data on how polypharmacy affects medication adherence in this population remain scarce. Most existing evidence comes from high-income countries or other African settings such as Ghana and may not reflect the realities of the Tanzanian healthcare system, where chronic care clinic capacity, medication supply chains, and health insurance coverage differ substantially. A clearer understanding of the polypharmacy– adherence relationship in the Tanzanian context is therefore essential to inform locally relevant clinical and policy interventions. This study, aimed at assessing the prevalence of polypharmacy and its association with medication adherence among outpatients with comorbid T2DM and hypertension attending Muhimbili National Hospital in Tanzania.

## Methodology

### Study design and setting

A cross-sectional study was conducted from March to June 2024 at the Upanga and Mloganzila branches of Muhimbili National Hospital (MNH), Dar es Salaam, Tanzania. These two branches were purposively selected because they host the principal diabetes and hypertension outpatient clinics at MNH, which is Tanzania’s national referral hospital, a teaching hospital, and research centre with 1,500 inpatient beds and approximately 2,000 outpatient visits per day. Data collection procedures, instruments, and interviewer training were identical at both sites. As a tertiary referral institution, MNH serves a patient population that may differ from those at district or primary care facilities, and this should be considered when interpreting the generalizability of findings.

### Study population

The study population comprised of outpatients diagnosed with T2DM and comorbid hypertension attending diabetes clinic at MNH. Eligible participants were outpatients aged 18 years or older with clinician-confirmed diagnoses of T2DM and hypertension documented in clinic medical records, who were actively receiving pharmacological treatment for both conditions. Patients with type 1 diabetes, gestational diabetes, or who demonstrated significant cognitive impairment, that is the inability to follow simple verbal instructions or provide coherent responses during the initial screening interaction were excluded. Written informed consent was obtained from all participants before enrollment, and participation was entirely voluntary..

### Sample size

The sample size was estimated using the formula n = Z²P(1−P)/d², where Z = 1.96 (95% confidence level), P = 0.368 (adherence prevalence from Osei et al., 2020), and d = 0.05 (margin of error), yielding a minimum of 360, inflated to 396 after applying a 10% non-response adjustment. The use of a Ghanaian prevalence estimate was necessitated by the absence of comparable published Tanzanian data and is acknowledged as a potential limitation. Of 430 patients approached, 396 consented and completed the interview, yielding a response rate of 92.1%.

### Sampling technique

Participants were recruited using a consecutive sampling, a non-probabilistic approach in which all eligible patients attending the diabetes clinic on each scheduled clinic day were approached for inclusion. Recruitment took place on all regular clinic days throughout the study period (March - June 2024). While this approach is efficient and minimizes within-day selection, it carries an inherent risk of selection bias because it does not constitute a random sample of all patients registered at MNH. Patients who missed appointments or attended infrequently were therefore less likely to be enrolled.

### Data collection

Data were collected through in-person interviews utilizing a structured questionnaire designed to capture socio-demographic characteristics (age, gender, marital status, level of education, occupation, mode of payment for medications), medication history (number and list of medications, frequency of doses, and use of herbal medicine), clinical characteristics (presence of other comorbidities, systolic and diastolic blood pressure, glucose levels, and frequency of follow-up visits), and adherence behavior. To minimize interviewer, bias all data collectors were trained and instructed not to suggest correct or preferred responses and were not informed of the specific study hypotheses regarding polypharmacy and adherence.

Medication adherence was evaluated using the Medication Adherence Report Scale (MARS-5), which consists of five questions addressing “stopping,” “skipping,” “changing dosages,” “forgetting,” and “using medication less than prescribed.” Participants rated their frequency of these behaviors as “always,” “often,” “sometimes,” “rarely,” or “never,” with scores ranging from 1 point for “always” to 5 points for “never.” The MARS-5 questionnaire was administered in Swahili. The English-language instrument underwent forward translation by a bilingual health professional and back-translation by an independent bilingual translator; discrepancies were resolved by consensus. Formal psychometric validation of the Swahili MARS-5 in the Tanzanian context has not been published, and this is acknowledged as a limitation. Blood pressure and fasting blood glucose readings were extracted from medical records. Polypharmacy was defined operationally as the concurrent use of five or more prescribed medications, consistent with the WHO definition.

### Data analysis

Data were initially entered into Microsoft excel and then exported to the Statistical Package for Social Sciences (SPSS) version 23 for analysis. Descriptive statistics were performed for categorical variables, utilizing frequency counts and percentages. Continuous variables that followed a normal distribution were summarized using means and standard deviations, while non-normally distributed continuous data were represented by medians and interquartile ranges. The MARS-5 scores were reported as percentages and frequencies. The total score, ranging from 5 to 25 points, was calculated by summing the scores for each question. Participants scoring the maximum of 25 were classified as adherent; those scoring below 25 were classified as non-adherent. The prevalence of polypharmacy (defined as the concurrent use of five or more medications) was expressed as a percentage. To identify factors associated with medication adherence, univariate modified Poisson regression with robust standard errors was first performed for each explanatory variable; those with p < 0.2 were retained for the multivariable model. Results are expressed as crude prevalence ratios (cPR) and adjusted prevalence ratios (aPR) with 95% confidence intervals (CI). Statistical significance was set at p < 0.05.

### Ethical approval and consent to participate

Ethical approval was granted by the Muhimbili University of Health and Allied Sciences (MUHAS) Research and Ethics Committee (Certificate Ref. No. DA.282/298/01L/902). Permission to conduct the study at MNH was obtained from the hospital administration. Written informed consent was obtained from all participants prior to enrollment. Participation was voluntary and participants were free to withdraw at any time without consequence. Confidentiality was maintained by assigning each participant a unique study identification number; no personal identifiers were recorded on data collection instruments, and all study documents were stored in locked cabinets accessible only to the principal investigator.

## Results

### Social demographic characteristics

This study involved 396 participants, of whom 59.1% were female. 53.3% of participants were aged between 50 and 59 years (53.3%), with a mean age of 59 years. Majority were married (85.4%). Education levels varied, with most participants having completed primary education (44.9%). 28.3% were employed, while 22.0% were unemployed. Notably, 98.2% of participants had health insurance, which facilitated access to medications, with 95.1% receiving all prescribed drugs through their insurance plans **Table 1**.

**Table 1:** Socio-demographics of the respondent (N=396).

| Variable | Categories | Frequency(N) | Percent (N %) |
| --- | --- | --- | --- |
| **Age category** | < 50 years | 28 | 7.1 |
|  | 50 - 59 years | 211 | 53.3 |
|  | 60 - 69 years | 128 | 32.3 |
|  | > 70 years | 29 | 7.3 |
| **Mean age (Standard Deviation)** |  | 59 (6.88) |  |
| **Gender** | Male | 162 | 40.9 |
|  | Female | 234 | 59.1 |
| **Marital status** | Married | 338 | 85.4 |
|  | Unmarried | 58 | 14.6 |
| **Education level** | No formal education | 15 | 3.8 |
|  | Primary education | 178 | 44.9 |
|  | Secondary education | 78 | 19.7 |
|  | Tertiary education | 125 | 31.6 |
| **Occupation** | Businessman/woman | 102 | 25.8 |
|  | Employed | 112 | 28.3 |
|  | Unemployed | 87 | 22.0 |
|  | Retired | 73 | 18.4 |
|  | Others | 22 | 5.5 |
| **Mode of payment for medication** | Health insurance | 389 | 98.2 |
|  | Self-sponsored | 7 | 1.8 |
| **Get all the drug from insurance** | No | 19 | 4.9 |
|  | Yes | 370 | 95.1 |

### Medication history

Polypharmacy, defined as the concurrent use of five or more medications, was present in 71% of participants (n = 281). The median number of medications was six, with 80.8% taking their medications twice daily. A small proportion (4.3%) reported using herbal medicine, primarily for management of diabetes (76.5%). All participants demonstrated awareness on how to take their prescribed medications, highlighting a positive aspect of patient education among the study participants, **Table 2**.

**Table 1:** Medication history of the respondent (N=396).

| **Variable** | **Category** | **Frequency (N)** | **Percent (N %)** |
| --- | --- | --- | --- |
| **Number of medications** | < 5 Medication | 115 | 29.0 |
|  | ≥ 5 Medication | 281 | 71.0 |
| **Number of medications (Median (Inter Quartile Range))** |  |  | 6 (4, 7) |
| **Frequency of daily dose of the medicine** | Once | 64 | 16.2 |
|  | Three times | 12 | 3.0 |
|  | Twice | 320 | 80.8 |
| **Use of herbal medicine** | No | 379 | 95.7 |
|  | Yes | 17 | 4.3 |
| **Herbal medicine indication** | Diabetes mellitus | 13 | 76.5 |
|  | Hypertension | 4 | 23.5 |
| **Know how to take all medication prescribed** | Yes | 396 | 100.0 |
|  | No | 0 | 0 |

**Table 2:** Clinical characteristics of the respondent (N=396).

| Variable | Category | Frequency (n) | Percent (N %) |
| --- | --- | --- | --- |
| **Other comorbid** | No | 339 | 85.6 |
|  | Yes | 57 | 14.4 |
| **Peptic ulcer disease** | Yes | 27 | 47.4 |
| **Cardiac disease** | Yes | 16 | 28.1 |
| **Kidney disease** | Yes | 10 | 17.5 |
| **Asthma** | Yes | 7 | 12.3 |
| **Goiter** | Yes | 5 | 8.8 |
| **Systolic blood pressure (mmHg)** | Controlled (<140mmHg) | 249 | 62.9 |
| | Uncontrolled<br>($\geq 140$mmHg) | 147 | 37.1 |
| **Median Systolic blood pressure (IQR)** |  | 139 (129–147) |  |
| **Diastolic blood pressure (mmHg)** | Controlled<br>($< 90$mmHg) | 348 | 87.9 |
| | Uncontrolled<br>($\geq 90$mmHg) | 48 | 12.1 |
| **Median Diastolic blood pressure (IQR)** |  | 84 (80–89) |  |
| **Fasting blood glucose level (Mmol/L)** | Low ($< 5.6$mmol/L) | 127 | 32.1 |
|  | Moderate (5.6 - 6.9 mmol/L) | 108 | 27.3 |
| | High ($\geq 7.0$mmol/L) | 161 | 40.7 |
| **Median fasting blood glucose level (IQR)** |  | 6.3 (5.1–8.0) |  |

Most patients (57.3%) were prescribed losartan + hydrochlorothiazide combination for the management of hypertension, whereas 29.5% of participants were on metformin plus glimepiride for blood glucose control and atorvastatin (53.3%) primarily for cholesterol control **Figure 1a**.

**Figure 1a:**
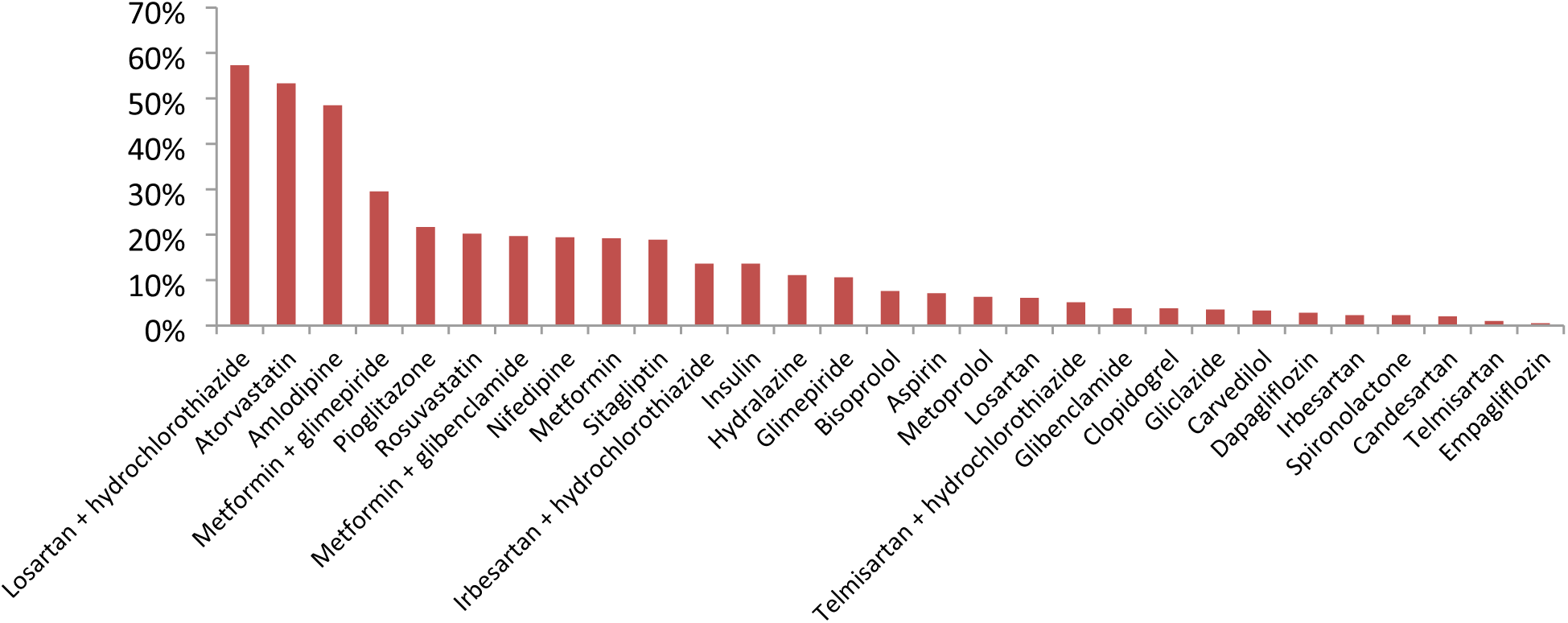
Antihypertensive and antidiabetic medications taken.

The most frequently taken supplements, **Figure 1b**, were diabetic support tablets (84.9%), NAT B fort (34%) and meloxicam (29%). Other drugs commonly taken by these patients included pregabalin (47.3%), esomeprazole (24%) and pantoprazole (23%), **Figures 1c**.

**Figure 1b:**
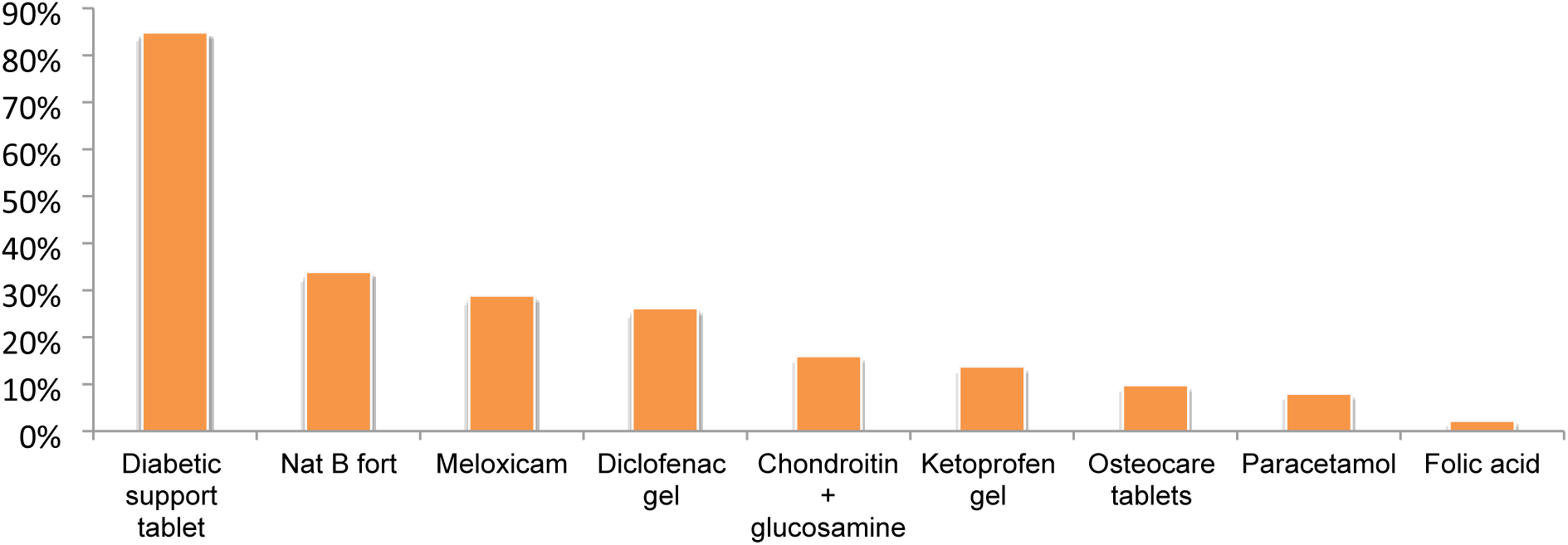
Pain relievers and supplements taken.

**Figure 1c:**
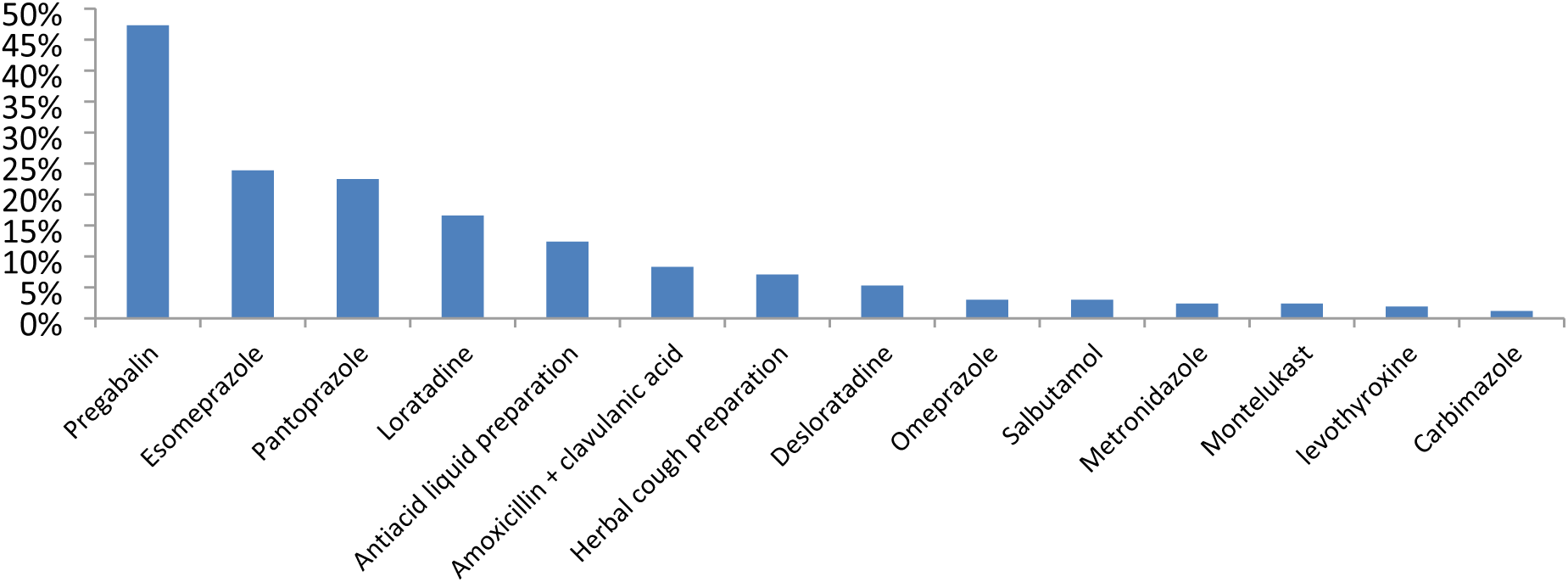
Antibiotics and other medications for additional comorbid taken.

### Clinical characteristics of study participants

Clinical assessments indicated that 14.4% of participants had additional comorbidities, including peptic ulcers (47%), kidney disease (28%), and heart disorders (17.5%). Blood pressure control was relatively good, with 62.9% of participants maintaining systolic blood pressure <140 mmHg and 87.9% maintaining diastolic blood pressure <90 mmHg. However, fasting blood glucose levels showed that 40.7% of participants had elevated glucose levels, indicating a need for better glycemic control, **Table 3**.

### Adherence behavior

Although the vast majority of participants reported never altering their dose or taking less medication than prescribed, a meaningful proportion acknowledged forgetting (32.3%) or deciding to miss a dose (16.7%), and these behaviors alone were sufficient to lower the total MARS-5 score below the adherence threshold of 25, accounting for the overall non-adherence prevalence of 44.9%.

### Factors associated with medication adherence

Multivariate analysis identified several factors, **Table 5** associated with medication adherence among patients with comorbid T2DM and hypertension. Herbal medicine use was independently associated with a lower probability of adherence; participants using herbal medicines were 28% less likely to be classified as adherent compared with non-users (aPR: 0.72, 95% CI: 0.64–0.82, p < 0.001). Polypharmacy was also associated with a lower probability of adherence; participants on five or more medications were 10% less likely to be adherent than those on fewer medications (aPR: 0.90, 95% CI: 0.81–0.99, p = 0.038). **(Table 5).**

**Table 4:** Participant responses on the medication adherence report (MARS-5) scale (N=396).

| **Adherence scale** | **Always**<br>**n (%)** | **Often**<br>**n (%)** | **Sometimes**<br>**n (%)** | **Rarely**<br>**n (%)** | **Never**<br>**n (%)** |
| --- | --- | --- | --- | --- | --- |
| I forgot to take my medicine | 1 (0.3) | 2(0.5) | 12(3.0) | 113(28.5) | 268(67.7) |
| I alter the dose of my medicine | 0 (0.0) | 0(0.0) | 1(0.3) | 2(0.5) | 393(99.2) |
| I stopped taking my medicine for a while | 0 (0.0) | 0(0.0) | 2(0.0) | 43(10.9) | 351(88.6) |
| I decided to miss out on the dose of my medicine | 0 (0.0) | 5(1.3) | 0(0.0) | 61(15.4) | 330(83) |
| I take less medicine than instructed | 0(0.0) | 0(0.0) | 0(0.0) | 1(0.3) | 395(99.7) |

**Table 5:**
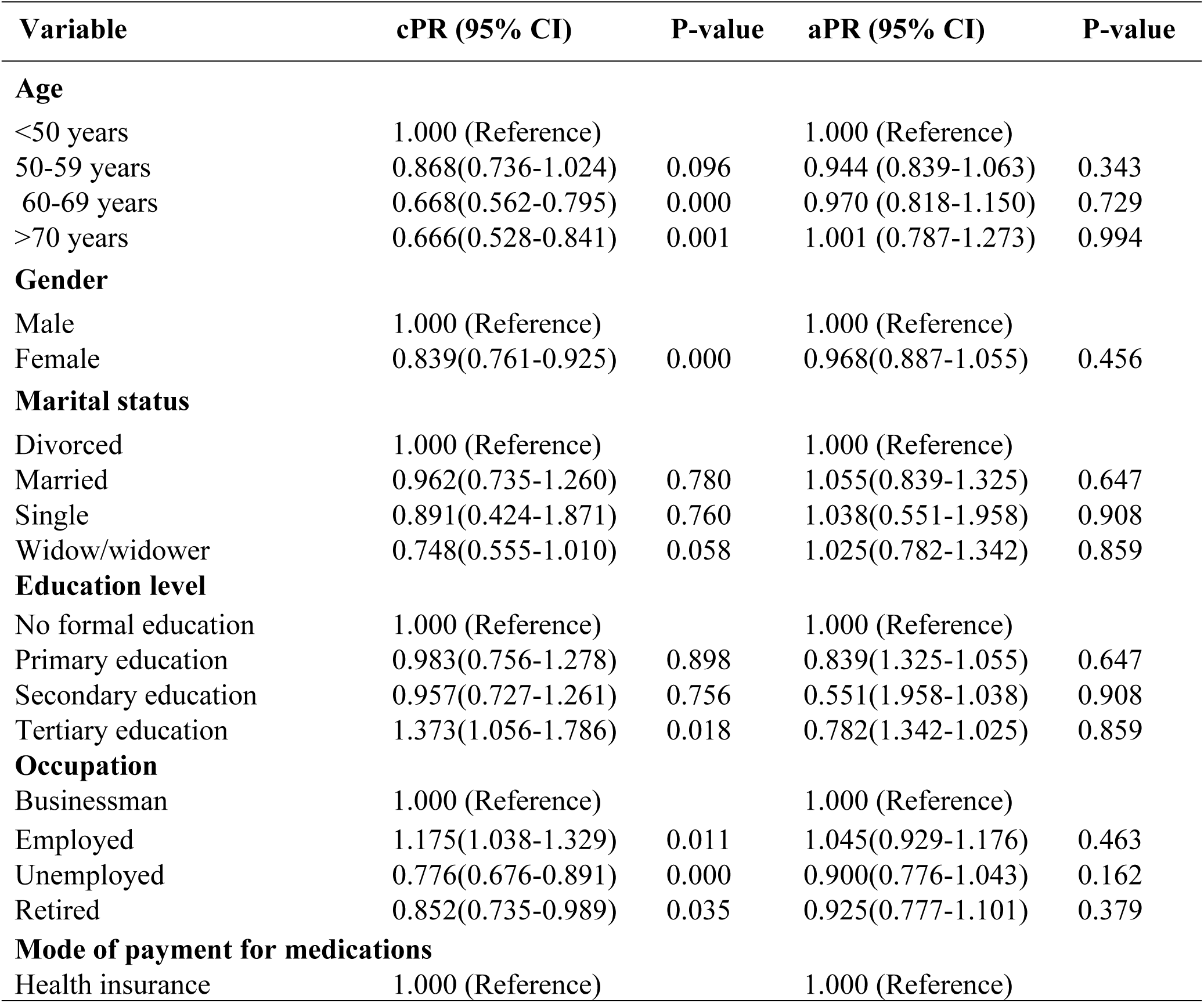
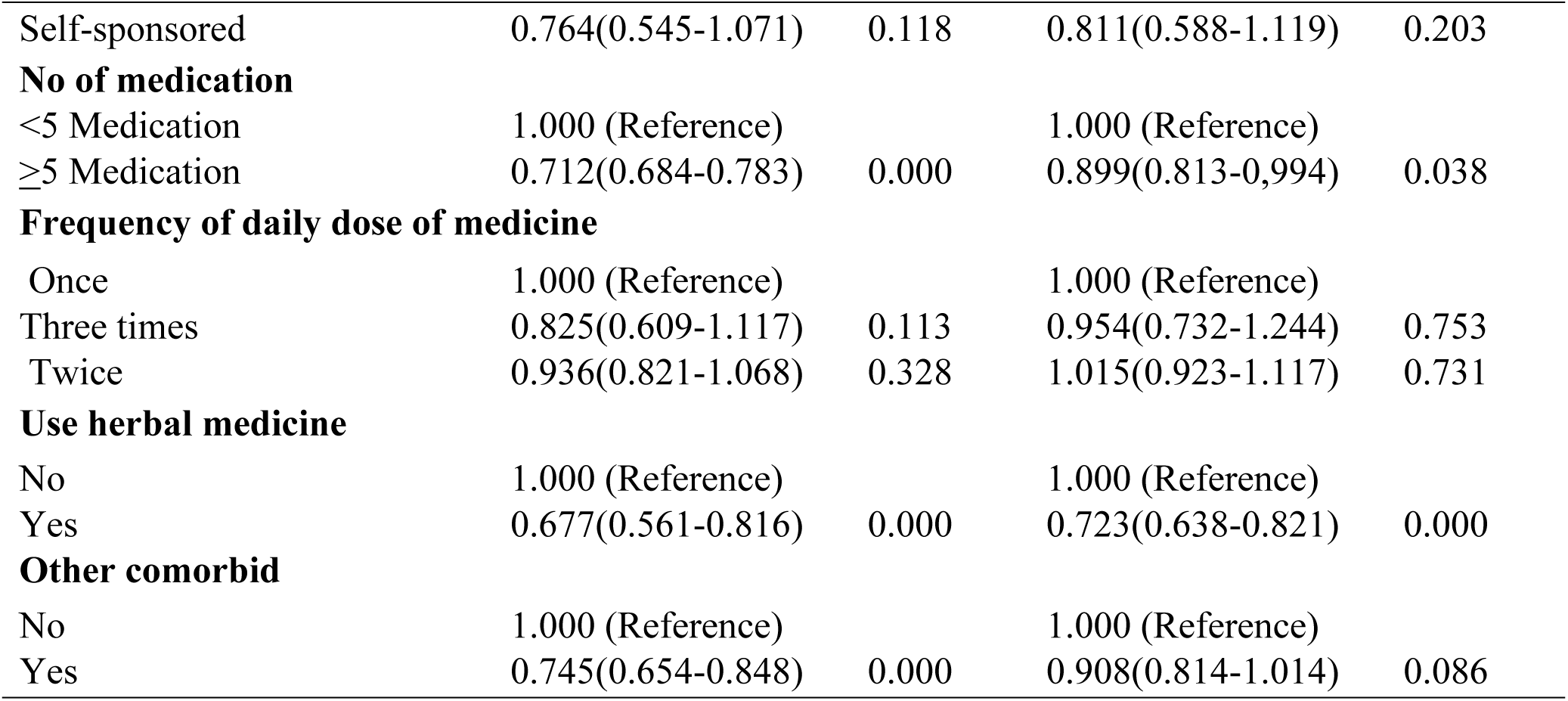
Modified Poisson regression model of factors associated with medication adherence (N=396).

Medication adherence was significantly associated with blood pressure control, whereby 79.8% of adherent participants had controlled systolic blood pressure versus 42.1%, and 95% of adherent participants had controlled diastolic blood pressure (**Figure 2**).

**Figure 2:**
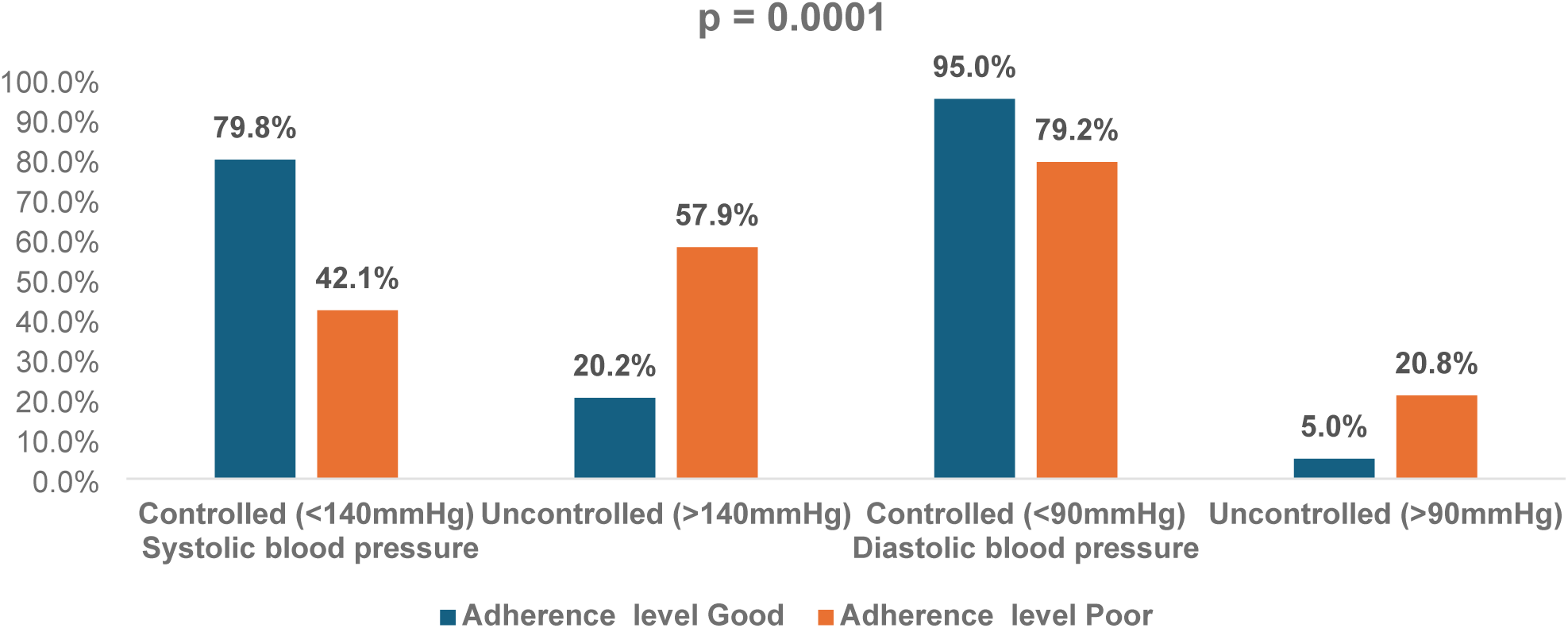
Association between adherence and blood pressure control.

A significant association was observed in blood pressure control whereby 79.8% of adherent participants had low to moderate fasting blood glucose levels compared to 34% of participants with poor adherence **(Figure 3).**

**Figure 3:**
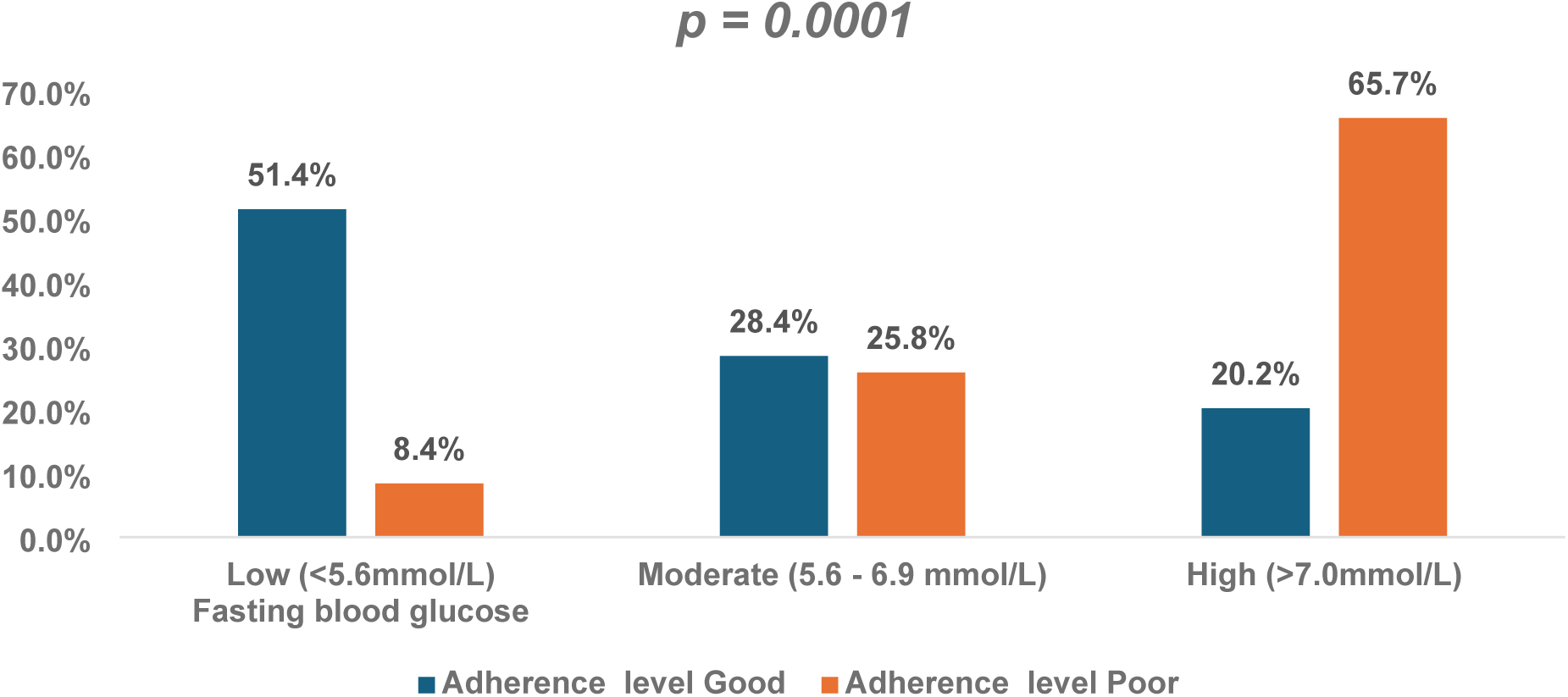
Association between adherence and fasting blood glucose control.

## Discussion

This study aimed at assessing the impact of polypharmacy on medication adherence in patients with comorbid type 2 diabetes and hypertension in Tanzania. Study findings suggest a significant prevalence of polypharmacy among this patient population. Notably, 71% of participants reported experiencing polypharmacy, indicating a high prevalence of concurrent medication use among individuals with multiple chronic conditions. This is consistent with findings from a study in Ghana (9), where 64.8% of participants with comorbid T2DM and hypertension reported polypharmacy, highlighting a similar trend in this patient population.

It is important to note that polypharmacy in patients with comorbid T2DM and hypertension is frequently clinically appropriate, given the need to control blood pressure, glucose, lipids, and associated cardiovascular risk concurrently. The clinical concern arises not from the number of medications, but from whether the regimen is regularly reviewed, rationalized, and tailored to individual patient needs. Furthermore, although the association between polypharmacy and lower adherence was statistically significant, the effect size was modest, and this finding should be interpreted with appropriate caution rather than taken as evidence that polypharmacy invariably undermines adherence.

Furthermore, the prevalence of medication adherence in this study was found to be 55.1%. This proportion is notably higher than the 36.8% medication adherence reported in previous studies utilizing the Medication Adherence Rating Scale (MARS-5) with similar cut-off values. This discrepancy may be attributed to geographic and demographic variations, as well as differences in healthcare access and patient education regarding the importance of adherence to treatment regimens. For instance, a study in India (10) found that better awareness and understanding of potential complications associated with diabetes and hypertension significantly contributed to higher adherence rates. The relatively higher adherence observed in this study may also partly reflect the near-universal health insurance coverage in our sample (98.2%), which facilitates consistent medication access and reduces the financial barrier to refilling prescriptions. However, caution is warranted: the MARS-5 relies on participants’ self-assessment of their behaviors and is susceptible to social desirability bias, which may lead to overestimation of true medication adherence.

Women constituted the majority of participants in this study (59.1%), which may reflect patterns of healthcare-seeking behavior and clinic attendance at this tertiary facility rather than any inherent difference in disease risk between sexes(11). Although women represented the larger group in bivariate analysis, gender was not independently associated with medication adherence in the multivariable model, and we therefore draw no conclusions about differential adherence by sex. The predominance of older participants (mean age 59 years) is consistent with the well-established increase in the prevalence of both T2DM and hypertension with advancing age (12)

The study also found a statistically significant association between polypharmacy and lower levels of medication adherence. The challenges associated with managing multiple medications can lead to increased rates of non-adherence, as patients may struggle to keep track of their medications and their associated schedules(13, 14). However, this relationship has not been consistently observed in other research, where some studies found no significant correlation between polypharmacy and medication adherence.

Interestingly, the use of herbal medicine was also linked to lower medication adherence in this study. A significant number of participants reported using herbal remedies without informing their healthcare providers. This finding aligns with previous studies that indicate the use of herbal medicines can negatively impact adherence to conventional treatment regimens for hypertension and diabetes (15). The current study did not distinguish between self-initiated and provider-recommended herbal medicine use; however, evidence from the broader East African context suggests that herbal use for chronic diseases is predominantly self-initiated and frequently undisclosed to prescribers (16)). In Tanzania, traditional medicine is culturally embedded and widely used alongside conventional care, making non-disclosure both common and clinically consequential, as it may lead to herb-drug interactions and scheduling conflicts that disrupt adherence to prescribed regimens(17, 18). Routine inquiry about herbal medicine use should therefore be integrated into all clinic consultations for patients with chronic conditions.

The study also identified a significant association between lower medication adherence and uncontrolled blood pressure, elevated fasting blood glucose levels. Inadequate adherence to prescribed medications often associated with unmet therapeutic goals, which is consistent with findings from other studies demonstrating that poor medication adherence is linked to uncontrolled hypertension (19) and poor glycemic control. Given the cross-sectional design of this study, causality cannot be inferred, and this relationship is likely bidirectional.

This study has several limitations that should be borne in mind when interpreting the findings. The cross-sectional design precludes causal inference; observed associations may reflect reverse causality or the influence of unmeasured confounders. The study was conducted at a single national tertiary referral hospital, which constrains generalizability to patients attending primary or district-level facilities in Tanzania. Medication adherence was assessed solely by self-report using the MARS-5; this method is susceptible to social desirability and recall bias, and objective measures such as pill counts, pharmacy refill records, or drug-level monitoring were not available. The use of consecutive sampling, while practical, is non-probabilistic and may under-represent patients with very poor adherence who attend clinic infrequently. The MARS-5 has not been formally validated in the Tanzanian context, and the strict maximum-score cut-off of 25 may overestimate non-adherence compared with studies that apply lower thresholds. Finally, the Ghanaian prevalence estimate used in the sample size calculation may not accurately reflect adherence rates in Tanzania, introducing uncertainty in the power assumptions.

## Conclusion

Healthcare professionals should implement educational initiatives that emphasize the importance of medication adherence and its direct impact on managing T2DM and hypertension. Addressing patients’ concerns regarding polypharmacy through regular counseling sessions is also crucial. To minimize polypharmacy, routine medication reviews should be conducted to assess the necessity of each prescribed medication. Simplifying medication schedules may enhance adherence and reduce the risk of drug interactions. Establishing a robust follow-up plan, including frequent check-ins and medication reminders, could further improve adherence.

Finally, it is essential for physicians, pharmacists, and nurses to collaborate as a multidisciplinary team to develop individualized treatment plans that account for the complexities of polypharmacy and its relationship with medication adherence. Routine medication reviews to assess and rationalize polypharmacy, combined with structured patient counselling on adherence, are recommended for integration into the standard management of patients with comorbid T2DM and hypertension at referral hospitals in Tanzania. Further multicenter research across different levels of the healthcare system is needed to determine whether these findings generalize beyond tertiary care settings.

## Declaration

### Availability of data and materials

The data sets used and analyzed during the current study are available upon reasonable request from the corresponding author.

### Authors’ Contribution

FBK: Conceptualization and design of research, data collection, data analysis, interpretation of the results, and manuscript writing. HI: Manuscript review and data interpretation, MAM: Manuscript review and data interpretation, AIM: Approved the research idea and research design, interpretation of the results, review of the manuscript, final approval of the manuscript. RFM: Approved the research idea and research design, interpretation of the results, review of the manuscript, final approval of the manuscript.

## Data Availability

All relevant data are within the manuscript and its Supporting Information files.

## Acknowledgments

We would like to extend our gratitude to the Muhimbili National Hospital (MNH) for granting permission and providing support to conduct this study. We also appreciate the contributions of the Muhimbili University of Health and Allied Sciences (MUHAS) for their guidance throughout this research. Finally, we acknowledge the patients who participated in this study, without whom this research would not have been possible.

## List of Abbreviations

DBP: Diastolic Blood Pressure
IDF: International Diabetes Federation
LMICs: Low- and Middle-Income Countries
MARS: Medication Adherence Reporting Scale
SBP: Systolic Blood Pressure
T2DM: Type 2 Diabetes Mellitus
WHO: World Health Organization

